# Higher pre-treatment evoked alpha-band oscillatory brain dynamics predict chronic pain reduction of non-invasive brain stimulation of non-motor targets

**DOI:** 10.64898/2025.12.31.25343260

**Authors:** Enrico De Martino, Margit Midtgaard Bach, Bruno Nascimento Couto, Anne Jakobsen, Stian Ingemann-Molden, Adenauer Casali, Thomas Graven-Nielsen, Daniel Ciampi de Andrade

## Abstract

Repetitive transcranial magnetic stimulation (rTMS) of non-motor cortical targets, including the left dorsolateral prefrontal cortex (DLPFC), anterior cingulate cortex (ACC), and posterosuperior insula (PSI), has been proposed as a treatment for chronic pain with variable clinical outcomes. Pre-treatment local cortical dynamics were hypothesized to serve as markers of chronic pain reduction. In this secondary analysis of a large clinical trial comparing different rTMS targets for pain relief, it was examined whether pre-treatment evoked cortical responses measured by electroencephalography after TMS of DLPFC, ACC, or PSI were associated with at least 30 percent reduction in pain intensity scored on a visual analogue scale. Forty-five patients with chronic pain received 12 sessions over eight weeks of 10 Hz rTMS to DLPFC, ACC or PSI. Cortical reactivity was quantified using global and local mean field power, and oscillatory dynamics were assessed using event-related spectral perturbation (ERSP) and inter-trial coherence (ITC) in the alpha-band (8–12 Hz). Responders (20 of 45, 44%) compared with non-responders showed higher pre-treatment alpha-band ERSP and ITC over the stimulated cortical targets (both P<0.05), and higher alpha-band ERSP and ITC values were negatively correlated with the percentage change in pain intensity (both P < 0.05). These results suggest that elevated pre-treatment TMS-evoked alpha-band oscillatory activity may indicate a higher probability of pain reduction to non-motor rTMS in chronic pain. This supports the development of enrichment strategies using cortical neurophysiology-based markers in neuromodulation trials aimed at individualized, precision-oriented treatments.

## INTRODUCTION

High-frequency repetitive transcranial magnetic stimulation (rTMS) of the primary motor cortex (M1) produces therapeutic benefit in people with chronic pain [1–4] and is included in guidelines and recommendations for the management of chronic pain [5]. It engages top-down pain modulatory mechanisms [6,7], changes cortical connectivity within pain-related networks [8,9], and restores abnormal intracortical excitability in chronic pain patients [10]. However, approximately half of the chronic patients show no meaningful pain relief from M1 rTMS. Among several strategies aiming at improving outcomes of pain neuromodulatory approaches [11], non-motor cortical targets have been increasingly investigated. The analgesic effects related to non-motor cortical rTMS may have different mechanisms of action than those of M1 rTMS [12–14], and possibly different effects on pain-related comorbidities, such as chronic pain-related mood symptoms [15,16].

The most reported non-motor rTMS targets are the left dorsolateral prefrontal cortex (DLPFC) [17,18], posterior superior insula (PSI) [19,20], and anterior cingulate cortex (ACC) [16,21]. Using rTMS of the left DLPFC has been investigated in several pain conditions, including migraine [22], burning mouth syndrome [23], and peripheral neuropathic pain [1]. The mechanisms of rTMS of the left DLPFC seem to involve top-down modulation of connections between the subgenual anterior cingulate cortex and amygdala, which are believed to act on mood and cognitive appraisal aspects of pain [24–26]. rTMS of the ACC has been investigated in patients with fibromyalgia [21] and in central neuropathic pain [16]. Neuroimaging evidence indicates that the ACC is involved in the cognitive and affective aspects of pain processing [27], which is in line with the anxiolytic effect of ACC rTMS [16]. There is increasing experimental and clinical evidence that neuromodulation strategies targeting the PSI can have analgesic effects and improve chronic pain [15,19,28]. Interestingly, PSI receives the largest number of corticospinal projections in primates [29]. Intracranial stimulation in humans has demonstrated that PSI modulates nociceptive pain [30] and can improve peripheral neuropathic pain [15]. Likewise, M1 rTMS, these non-motor rTMS targets have demonstrated analgesic effects in people with chronic pain, but with highly variable response rates across studies. Identifying neurophysiological biomarkers that predict the most effective non-motor cortical target for pain relief could enable personalized treatment approaches.

Using a neurophysiological approach that combines TMS with electroencephalography (TMS–EEG), a method used to assess cortical function [31], we have shown that baseline neurophysiological markers of cortical reactivity and oscillatory dynamics predict a meaningful analgesic effect of M1 rTMS in people with chronic pain [9]. Specifically, patients with lower global and local mean-field power, along with reduced alpha-band power and synchronization, experienced greater pain relief [9]. These findings suggest that neurophysiological measures obtained with TMS-EEG can prospectively identify those most likely to benefit from M1 rTMS therapy [32,33]. Here, we tested whether responders to rTMS delivered to non-motor cortical targets (DLPFC, ACC, and PSI) exhibit lower pre-treatment global and local mean field power, along with reduced alpha-band power and synchronization, compared with non-responders, mirroring the signature we previously observed in M1.

## METHODS

### Participants

This was a secondary, pre-planned exploratory analysis from a large randomized controlled trial assessing the efficacy of TMS-EEG-informed selection of the most appropriate target to be used for analgesic rTMS (ClinicalTrials.gov NCT06395649, [34]). The present study included all 45 patients randomized to non-motor targets (DLPFC, n = 16; ACC, n = 10; PSI, n = 19). Eligible participants were adults between 18 and 80 years with chronic pain lasting ≥6 months. The trial included patients with neuropathic, nociplastic, and nociceptive pain mechanisms, including both primary and secondary pain conditions, provided that baseline pain intensity was at least moderate (numerical rating scale (NRS) > 3/10). Exclusion criteria were current major psychiatric disorders, substance use disorder, unstable medical conditions, and standard contraindications to rTMS (e.g., severe head trauma, prior neurosurgery, current or past epilepsy, intracranial hypertension, or implanted ferromagnetic/electrical devices). Additional exclusions were pregnancy or breastfeeding, and inability to complete study follow-up. Furthermore, results from a control group of 20 healthy, pain-free participants (age: 48 ± 13 years; height: 173 ± 13 cm; weight: 76 ± 13 kg; 13 females) were used to provide reference ranges for TMS–EEG metrics. Each control participant underwent stimulation at all three cortical targets (DLPFC, ACC, and PSI), yielding a total of 60 TMS–EEG recordings used to derive reference values. The protocol was approved by the local Ethics Committee (Den Videnskabsetiske Komité for Region Nordjylland: N-20230076 and N-20230040 for the healthy participants) and conducted in accordance with the Declaration of Helsinki. All participants received an oral introduction to the study and provided written informed consent before any study procedures.

### Study design

Prior to rTMS, patients underwent TMS-EEG assessments. The rTMS intervention began the following week and spanned 8 weeks, consisting of an induction phase of daily sessions for 5 consecutive days, followed by a maintenance phase with 1 session per week for 7 weeks (12 sessions in total).

Responder status was defined based on a ≥30% reduction in mean pain intensity from baseline (the average pain during the week preceding the TMS–EEG session) to the final day of therapy (asked as how is your pain ‘right now’). Pain intensity was measured on a 0–100 visual analogue scale (VAS), where 0 indicates “no pain” and 100 “worst pain imaginable” [35].

For ACC and PSI, the TMS-EEG and rTMS were performed contralaterally to the side of the most intense pain. If no dominant pain side was reported, stimulation was applied to the left hemisphere [1]. For the DLPFC, the TMS-EEG and rTMS were always performed on the left hemisphere since the analgesic effects were reported only from the left site [36–38].

In the health control group, participants underwent a single TMS-EEG session at all three cortical targets (DLPFC, ACC, and PSI). The sequence of assessment targets was randomized across all participants.

### Clinical outcomes

At baseline and week 8 (final day of the treatment), participants also completed questionnaires including: (i) average pain VAS score over the past 7 days, (ii) pain VAS score in the past 24 hours, (iii) the Brief Pain Inventory–Short Form (BPI), (iv) a pain body map, (v) current analgesic medications, (vi) sleep quality, (vii) fatigue level, (viii) the Hospital Anxiety and Depression Scale (HADS), (ix) health-related quality of life (EQ-5D) and (x) Patient Global Impression of Change (PGIC).

From the BPI [39], pain severity was calculated as the mean of the four intensity items (worst, least, average, current), and pain interference as the mean of seven interference items (general activity, mood, walking ability, normal work, relations with others, sleep, enjoyment of life). The number of painful body regions was computed from the body map to index the spatial distribution of pain [40].

Sleep quality and fatigue levels were also assessed using VAS. For sleep, participants rated the question ‘How did you sleep last night?’ on a scale from 0 (bad sleep) to 100 (good sleep). For fatigue, they rated the question ‘How tired have you been in the last 24 hours?’ on a scale from 0 (no tiredness) to 100 (worst imaginable tiredness).

HADS is a 14-item self-report questionnaire designed to investigate symptoms of anxiety and depression, with higher scores indicating greater symptom severity [41].

The EQ-5D assesses mobility, self-care, usual activities, pain/discomfort, and anxiety/depression, together with a health-state VAS [42].

The PGIC is a 7-point scale of overall improvement (“very much worse” to “very much improved”) capturing perceived change since baseline [43].

### Electroencephalographic recordings during transcranial magnetic stimulation

An electroencephalogram was recorded with 64-channel passive electrodes (EasyCap) arranged according to the 10–5 system, with Cz at the vertex, using a TMS-compatible amplifier (g.HIamp, g.tec medical engineering GmbH). The ground electrode was placed on the right zygoma, and the online reference on the right mastoid. Horizontal eye movements were monitored with two electrodes positioned laterally to the eyes. Electrode impedance was maintained below 5 kΩ. Signals were sampled at 4800 Hz. TMS pulses were delivered with a biphasic stimulator (MagPro R30, MagVenture A/S, Farum, Denmark) using a figure-of-eight shaped coil (Cool-B35) for DLPFC stimulation, whereas a double-cone coil was used for ACC and PSI (Cool-D-B80) to ensure adequate depth of penetration [16,44].

During TMS-EEG recordings, patients were seated comfortably in an ergonomic chair and instructed to maintain a relaxed gaze on a fixation point to minimize oculomotor artifacts. To attenuate auditory responses to the TMS click sound, continuous noise masking was applied using the TMS Auditory Artefact Control (TAAC) toolbox [45], with noise-cancellation in-ear headphones (ER3C Etymotic 50 Ohm). Two elastic meshes and a plastic stretch film (GVB-geliMED GmbH) were applied over the EEG cap to minimize electrode movement and somatosensory artefacts induced by coil contact.

A neuronavigation system (InVesalius Navigator, 3.1 with a Polaris Vicra, NDI, Ontario, Canada) was used for target identification and monitoring during recordings. The DLPFC target was located in the middle frontal gyrus using the method described [46]. This location corresponds to the area below the F3 electrode in the 10–20 EEG system. The stimulation intensity was set to 110% of the resting motor threshold (rMT) of the right-hand muscles [17,47]. The M1 hot spot of the right-hand muscles was identified by locating the hand muscle representation in the left hemisphere at the largest observable hand twitch, and the rMT was defined as the lowest TMS intensity required to elicit a visible hand twitch [48].

The ACC target was defined as 4 cm anterior to the motor hotspot of the lower limb representation [49]. The M1 hotspot was defined as the coil position that elicited a visible leg twitch at a given stimulation intensity. The rMT of the leg muscle was determined as the lowest TMS intensity required to produce a visible leg twitch [16], nd TMS–EEG was performed at 90% of the leg rMT [50].

The PSI target was identified using a previously established and validated fast-PSI formula developed based on neuroanatomical landmarks and functional imaging studies [51]. This method first identifies the vertex, the intersection of the nasion-inion and tragus-tragus distances on the scalp. Next, the Nasion-PSI line is calculated by multiplying the nasion-inion distance by a correction factor. Similarly, the Vertex-PSI line is calculated by multiplying the vertex-tragus distance by another correction factor. The Fast-PSI target is then located at the intersection of these two lines [16,19]. The TMS-EEG stimulation intensity was set at 90% of the leg rMT [50].

A real-time visualization tool (rt-TEP) was used to ensure detectable TMS-evoked potentials (TEPs) in the three cortical targets [52]. This interface enabled adjustments to the TMS coil orientation and intensity across participants to minimize unwanted artifacts (e.g., muscle activity) and to achieve an early peak-to-peak amplitude (25–120 ms) between 6 μV and 25 μV, measured from the average of 30 trials at the EEG electrode closest to each target. These adjustments were necessary to guarantee a clear cortical response from the stimulated area while maximizing the signal-to-noise ratio in the EEG recordings [50].

The TMS-neuronavigation and rt-TEP were utilized throughout the study to monitor the TMS coil location. Approximately 200 pulses were administered per target, with interstimulus intervals jittered between 2000 and 2400 ms [53].

### Electroencephalographic preprocessing

EEG data were analyzed using Python (Python Software Foundation). The signals were segmented into trials of 1600 ms around the TMS pulse, which occurred at time zero (±800 ms). To remove TMS artifacts, the segment −15 to 0 ms was used to substitute the recordings from 0 to 15 ms in the DLPFC TMS-EEG epoch [33]. The same procedure was applied for ACC and PSI TMS targets, but with a larger peri-TMS interval (0-25 ms) due to the double-cone coil’s electric field [50]. Afterwards, epochs and channels containing isolated noise, eye blinks, eye movements, or muscle artifacts were identified and removed, resulting in a minimum of 150 trials per target [54]. Independent component analysis (fast ICA in MNE-Python [55]) was applied to the combined dataset to remove additional residual artifacts [56]. The EEG data were band-pass filtered (1-45 Hz, Butterworth, 3rd order), downsampled to 1200 Hz, and re-referenced to the average reference. Lastly, signals from any erroneous channels (e.g., disconnected or high-impedance) were interpolated based on neighboring channels using spherical splines [57].

### Metrics for cortical reactivity and oscillatory dynamics

TMS-evoked potentials from left DLPFC, left and right ACC, and left and right PSI are shown in Figure 1. Global mean field power (GMFP) was calculated to assess global cortical excitability, calculated as the root-mean-squared value of the TEP across all electrodes within the 15–120 ms interval following the TMS for DLPFC stimulation, and within the 20–120 ms interval for ACC and PSI stimulation [50]. To assess local cortical excitability, the local mean field power (LMFP) was calculated across electrodes near the TMS target [50].

**Figure 1:**
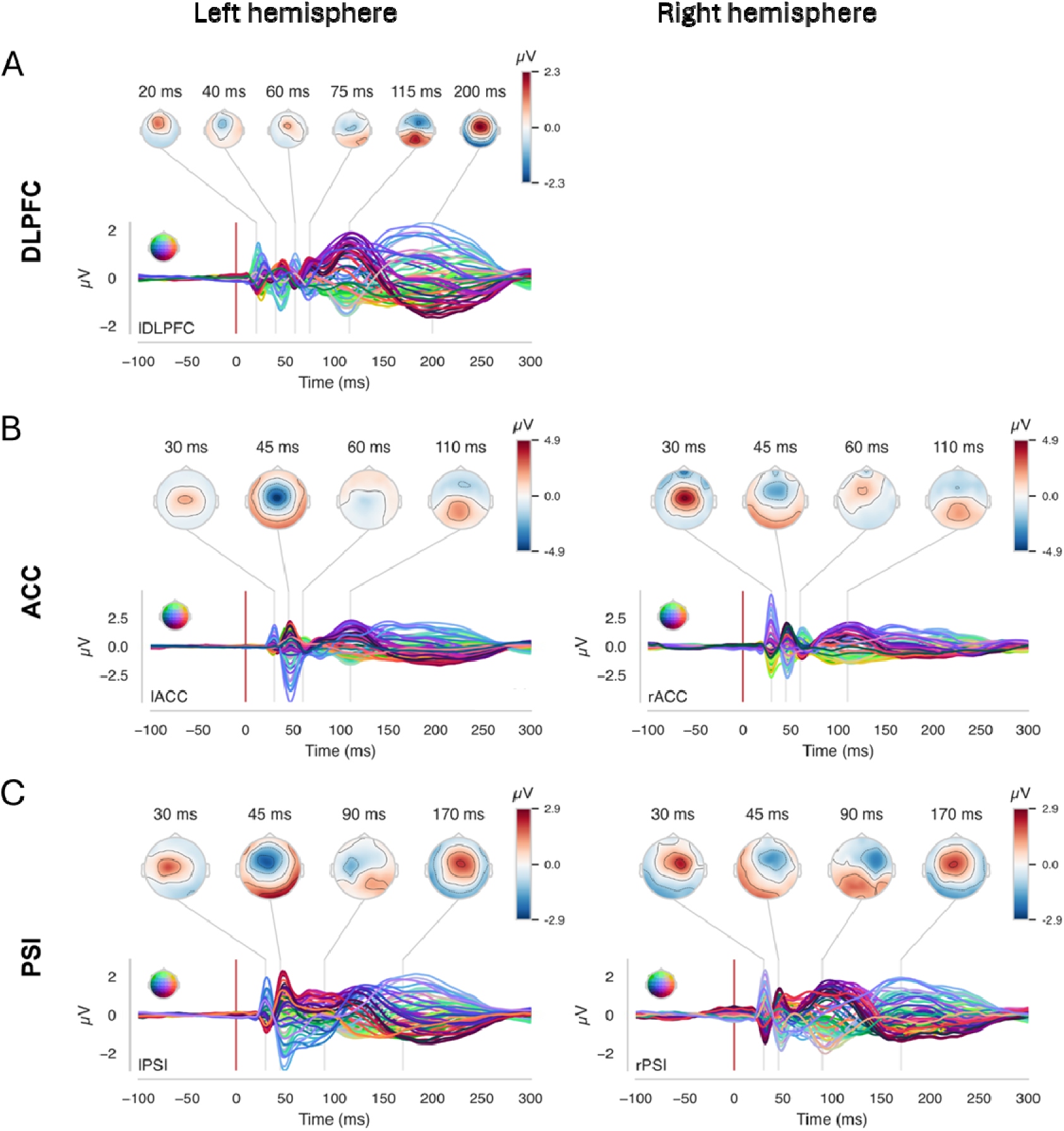
TMS-evoked potentials (TEPs) and corresponding scalp topographies for the three stimulation targets. **(A)** Grand-average TEP waveforms recorded across all participants are shown for the left DLPFC target. Representative scalp topographies at selected latencies illustrate the evolving spatial distribution of TMS-evoked activity. **(B)** Grand-average TEPs for right and left ACC, with corresponding topographies. **(C)** Grand-average TEPs for right and left PSI, with corresponding topographies.

For left DLPFC stimulation, LMFP was computed over the 15–120 ms post-TMS interval using electrodes F3, F1, FC3, and FC1. For ACC and PSI stimulation, LMFP was computed over the 25–120 ms interval. Left and right ACC responses were derived from electrodes FCz, Cz, FC1, FC2, C1, and C2. For left PSI, responses were derived from electrodes FC7, FC3, C7, C5, and C3, and for right PSI, from electrodes FC8, FC4, C8, C6, and C4.

To assess TMS-evoked oscillatory dynamics in the electrode cluster around the stimulated area, time–frequency maps were computed in the 8–45 Hz range using Morlet wavelets with 3.5 cycles [58]. The analysis focused on the alpha (8–12 Hz) band, and two parameters were extracted: ERSP and ITC. The selection of the alpha band was based on evidence showing that alpha activity is altered in chronic pain [59] and that alpha oscillatory dynamics during M1-rTMS predict the therapeutic efficacy of rTMS [9]. ERSP quantified changes in spectral power across frequency over trials. It was calculated as the average spectral power ratio of individual TMS-EEG trials relative to a pre-stimulus baseline (−600 to −50 ms), capturing how TMS-evoked oscillatory power evolves across frequency bands. Statistical significance was determined by comparing ERSP values to pre-stimulus baseline using bootstrapping (500 permutations, two-sided test, p < 0.05, with false discovery rate correction for multiple comparisons). Significant ERSP values were then averaged across time points and electrodes to characterize frequency-specific power modulation evoked by TMS [50,60].

ITC was extracted to evaluate the phase synchrony of cortical oscillatory activity across trials. ITC was computed by normalizing the complex-valued time–frequency data from each trial, averaging across trials, and taking the absolute value. Higher ITC values (close to 1) indicate stronger phase-locking and greater neural synchrony across trials. Significance was assessed using bootstrapping against baseline (500 permutations, one-sided test, p < 0.05, with false discovery rate correction for multiple comparisons). Significant ITC values were averaged across electrodes, time windows, and frequency bins [50,60].

### Treatment by repetitive transcranial magnetic stimulation

The treatment was delivered using a transcranial magnetic stimulator (MagPro R30; MagVenture A/S, Farum, Denmark) with a figure-of-eight Cool-B35 coil for the DLPFC, and a double-cone coil Cool-D-B80 for the ACC and PSI. For DLPFC and ACC, patients were seated comfortably in a reclining chair and instructed to remain as relaxed as possible throughout each session. For PSI stimulation, patients lay on a bed in a side-lying position, with the stimulated hemisphere oriented upward and accessible for treatment. Each rTMS session involved 30 trains of 100 TMS pulses delivered over 10 seconds (10 Hz), with 20-second pauses between trains, totaling 3,000 pulses over 15 minutes [1,33,50]. Stimulation intensity was set at 110% of the hand rMT for the DLPFC [17] and 90% of the TA rMT for the ACC and PSI [16]. All adverse events were recorded throughout the study period. During rTMS treatment, patients were allowed to continue their concurrent medications and other treatments.

### Statistical analysis

Statistical analyses were conducted using the Statistical Package for Social Sciences (SPSS, version 25; IBM, Chicago, IL). All results are presented as means and standard deviations. Statistical significance was set at a two-sided 5% level, with adjustment for multiple comparisons applied when appropriate. Normality of data distributions was assessed in the Responder and Non-responder groups using the Shapiro–Wilk test and visual inspection of histograms to identify extreme violations.

Separate two-way mixed-model analyses of variance (ANOVAs) were used to compare clinical outcomes, including average pain VAS in the last 24 hours, average pain VAS over the past 7 days, quality of life, BPI (pain severity and interference), HADS, and medication use. Time (pre-treatment, week 8) was included as the within-subject factor, and stimulation target (DLPFC, ACC, PSI) as the between-subject factor. Bonferroni-corrected pairwise comparisons with adjusted 95% confidence intervals (CIs) and P values were reported. Greenhouse–Geisser corrections were applied when the assumption of sphericity was violated, and effect sizes were quantified using partial eta-squared (η²partial).

To determine whether treatment response was dependent on stimulation target, a chi-square test of independence was conducted between responder status (Responder vs. Non-responder) and stimulation site (DLPFC, ACC, PSI). No significant association was observed (χ²(2) = 3.97, p = 0.138). Based on this result, stimulation targets were pooled for subsequent neurophysiological analyses comparing Responders and Non-responders.

For neurophysiological outcomes, Student’s t-tests were used for normally distributed data and Mann–Whitney U tests for non-normally distributed data to compare Responders and Non-responders. Each neurophysiological variable was evaluated relative to reference data obtained from 20 healthy control participants. For each measure, the median and 25th–75th percentile range were used to define the reference intervals shown in Figure 3. These reference data were provided for descriptive context only and were not included in any statistical analyses.

Spearman’s correlation analyses were conducted across all patients to examine associations between percentage pain VAS reduction and neurophysiological measures that showed between-group differences. Bonferroni correction was applied to account for multiple correlations.

## RESULTS

### Patient characteristics, treatment dose, compliance, and adverse events

The demographic, clinical characteristics, and main diagnosis are reported in Tables 1 and 2. Patients in the Responder and Non-responder groups had similar age and height. Of the 45 patients, 20 (44%) were classified as Responders to rTMS. Among females, 53% were classified as Responders. In contrast, 27% of males were classified as Responders.

**Table 1.** Demographic and clinical characteristics of Responders (N = 20) and Non-responders (N = 25).

|  | Responders | Non-responders |
| --- | --- | --- |
| Age (years) | 52±12 | 56±10 |
| Height (cm) | 173±10 | 174±8 |
| Weight (kg) | 84±12 | 94±20 |
| Male (N) | 4 | 11 |
| Female (N) | 16 | 14 |
| Nociceptive pain (N) | 5 | 6 |
| Neuropathic pain (N) | 9 | 9 |
| Nociplastic pain (N) | 6 | 9 |
| Primary pain type (N) | 6 | 9 |
| Secondary pain type (N) | 14 | 14 |
| Years with pain (years) | 13±11 | 11±11 |

**Table 2.**
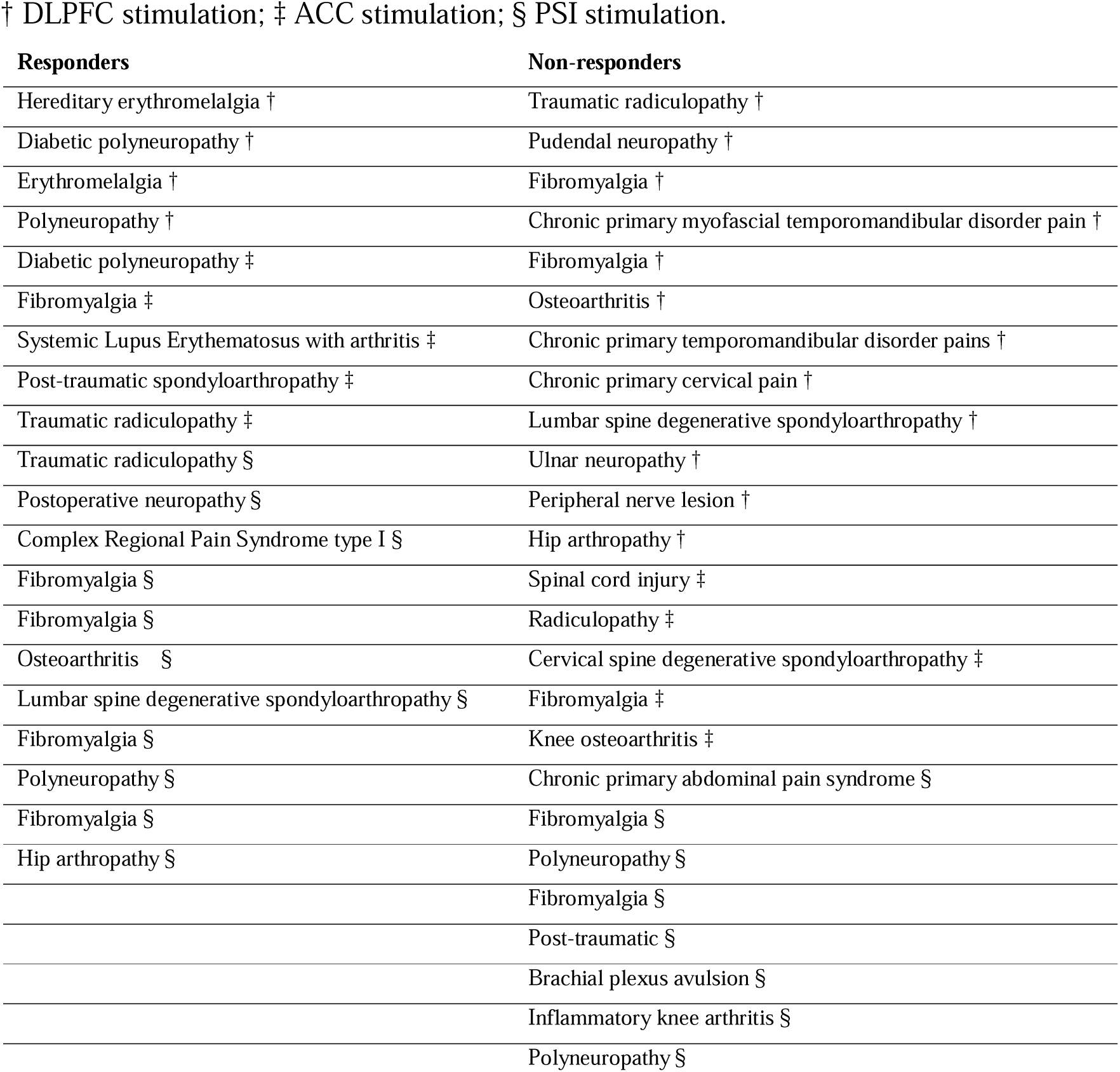
Main diagnosis.

| Responders | Non-responders |
| --- | --- |
| Hereditary erythromelalgia † | Traumatic radiculopathy † |
| Diabetic polyneuropathy † | Pudendal neuropathy † |
| Erythromelalgia † | Fibromyalgia † |
| Polyneuropathy † | Chronic primary myofascial temporomandibular disorder pain † |
| Diabetic polyneuropathy ‡ | Fibromyalgia † |
| Fibromyalgia ‡ | Osteoarthritis † |
| Systemic Lupus Erythematosus with arthritis ‡ | Chronic primary temporomandibular disorder pains † |
| Post-traumatic spondyloarthropathy ‡ | Chronic primary cervical pain † |
| Traumatic radiculopathy ‡ | Lumbar spine degenerative spondyloarthropathy † |
| Traumatic radiculopathy § | Ulnar neuropathy † |
| Postoperative neuropathy § | Peripheral nerve lesion † |
| Complex Regional Pain Syndrome type I § | Hip arthropathy † |
| Fibromyalgia § | Spinal cord injury ‡ |
| Fibromyalgia § | Radiculopathy ‡ |
| Osteoarthritis § | Cervical spine degenerative spondyloarthropathy ‡ |
| Lumbar spine degenerative spondyloarthropathy § | Fibromyalgia ‡ |
| Fibromyalgia § | Knee osteoarthritis ‡ |
| Polyneuropathy § | Chronic primary abdominal pain syndrome § |
| Fibromyalgia § | Fibromyalgia § |
| Hip arthropathy § | Polyneuropathy § |
|  | Fibromyalgia § |
|  | Post-traumatic § |
|  | Brachial plexus avulsion § |
|  | Inflammatory knee arthritis § |
|  | Polyneuropathy § |

Among the Responders, 11 received PSI stimulation, 5 received ACC stimulation, and 4 received DLPFC stimulation. Among the Non-Responders, 8 received PSI stimulation, 5 received ACC stimulation, and 12 received DLPFC stimulation.

Out of the 12 total rTMS sessions, the average number of attended therapeutic sessions was 11.5 ± 1.3 (96%) among Responders and 11.4 ± 1.6 (95%) among Non-responders. Two patients in the Non-responder group missed five and six consecutive sessions, respectively, and one patient in the Responder group missed two consecutive sessions. All other missed sessions were isolated.

There were no significant differences between the Responder and Non-responder groups in TMS intensity used during TMS–EEG (Table 3; all p > 0.05).

**Table 3.**
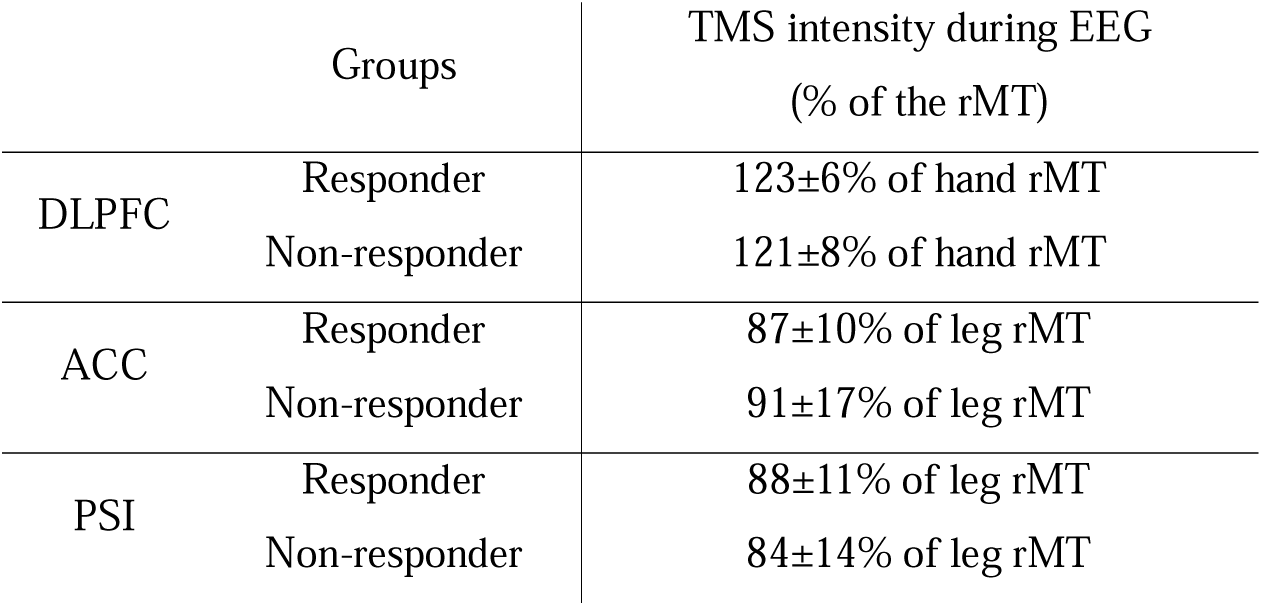
Mean ± standard deviation of transcranial magnetic stimulation (TMS) intensities used for TMS-evoked electroencephalographic (EEG) recordings, reported as a percentage of the resting motor threshold (rMT) of the hand or leg twitch.

Adverse events were reported in 30% of the rTMS sessions. They included “headache”(8%), “feel dizzy” (7%) ,“a trembling or numb sensation or the feeling of electricity in a part of the body” (6%), “twitching movements of an arm, leg or other body part” (5%), and “unusual smells, tastes, or emotions” (3%). “Unexplained confusion, drowsiness or weakness” and “Unusual experiences - out of body experiences, a feeling of being distracted, your body feels different” were reported in 1% of sessions. Approximately half (53%) of the side effects were reported during the 5 days of the induction week, and the remaining during the 7 weeks of maintenance.

### Clinical outcomes

The ANOVA revealed significant Time × Group interactions for average pain VAS intensity in the last 7 days (F_1,43_ = 17.025; P <0.001; η²_partial_ = 0.28), average pain VAS intensity in the last 24 hours (F_1,43_ = 9.480; P = 0.004; η²_partial_ = 0.18), and BPI pain severity (F_1,43_ = 8.177; P =0.007; η²_partial_ = 0.16). At the end of the treatment, pairwise comparisons indicated lower pain intensity in the last 7 days in Responders compared with Non-responders (P < 0.001; 95% CI [−34.6 −12.8]), the average pain VAS intensity in the last 24 hours (P < 0.001; 95% CI [−40.2 −16.3]), and BPI pain severity (P < 0.001; 95% CI [−2.9 −0.9]) (Figure 2).

**Figure 2:**
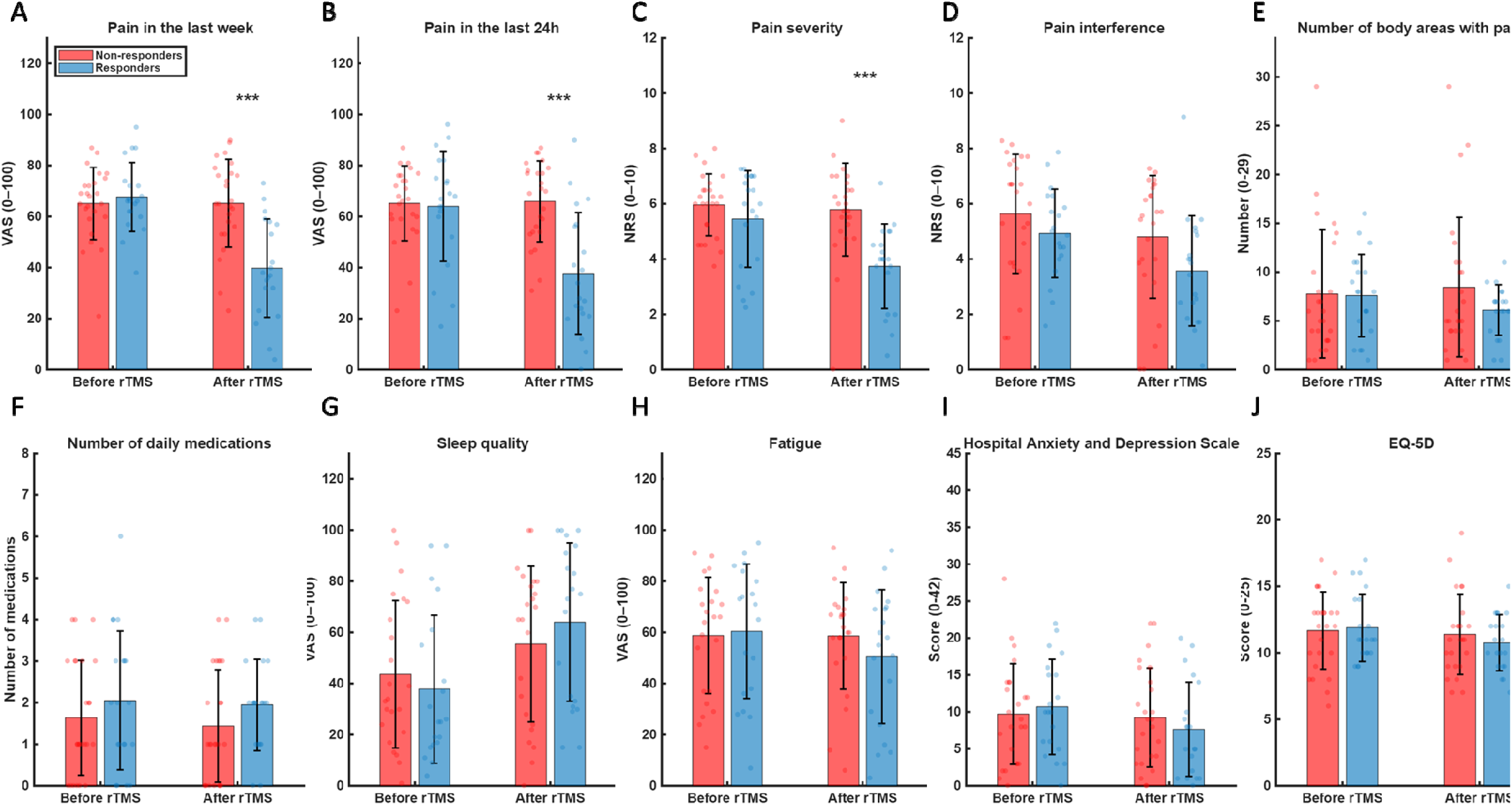
Secondary clinical measures assessed before and after rTMS treatment in Responder (blue) and Non-responder (red) groups. Bar plots display group means ± SD with individual data points overlaid. **(A)** Pain intensity in the last 24 h, **(B)** pain intensity in the last week, **(C)** BPI pain severity, **(D)** BPI pain interference, **(E)** number of body areas with pain, **(F)** number of medications, **(G)** sleep quality, **(H)** fatigue, **(I)** Hospital Anxiety and Depression Scale (HADS), and **(J)** EQ-5D quality-of-life score. Significant group differences are indicated (***p < 0.001).

**Figure 3:**
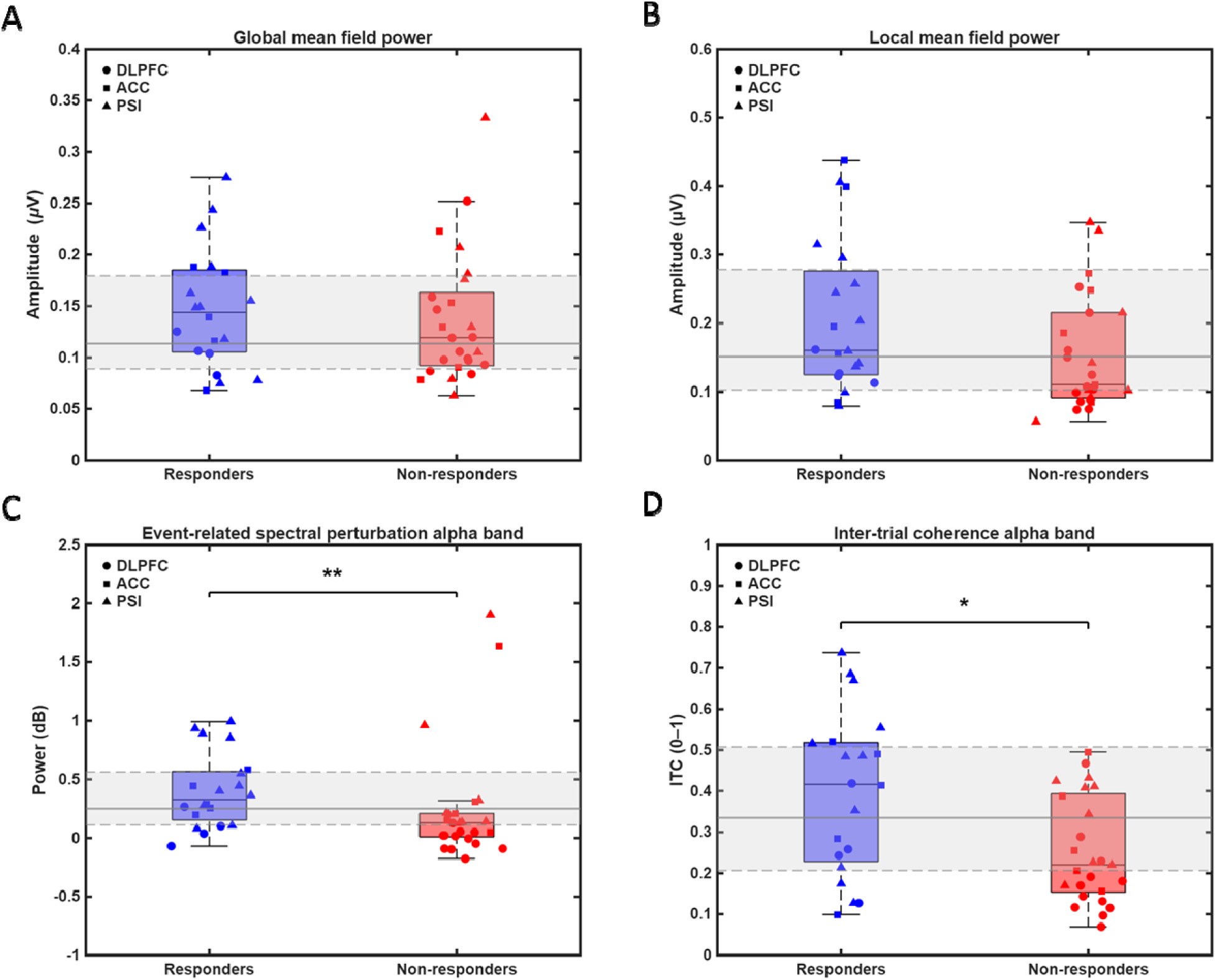
Neurophysiological outcomes in Responders (blue) and Non-responders (red) before rTMS treatment. Boxplots represent median ± quartiles, with individual participant data shown as dots. **A)** Global mean field power; **B)** Local mean field power; **C)** Event-related spectral perturbation in the alpha band; **B)** Inter-trial coherence in the alpha band. Asterisks indicate statistically significant differences between groups (*p < 0.05; **p<0.01).

The ANOVA did not reveal any interaction for BPI pain interference, the number of body areas, the number of daily medications, sleep quality, fatigue, HADS, and quality of life (all F_1,43_ ≥ 3.571; P ≥ 0.066; η²_partial_ ≥ 0.08). The list of daily medications is reported in Table 4.

**Table 4.**
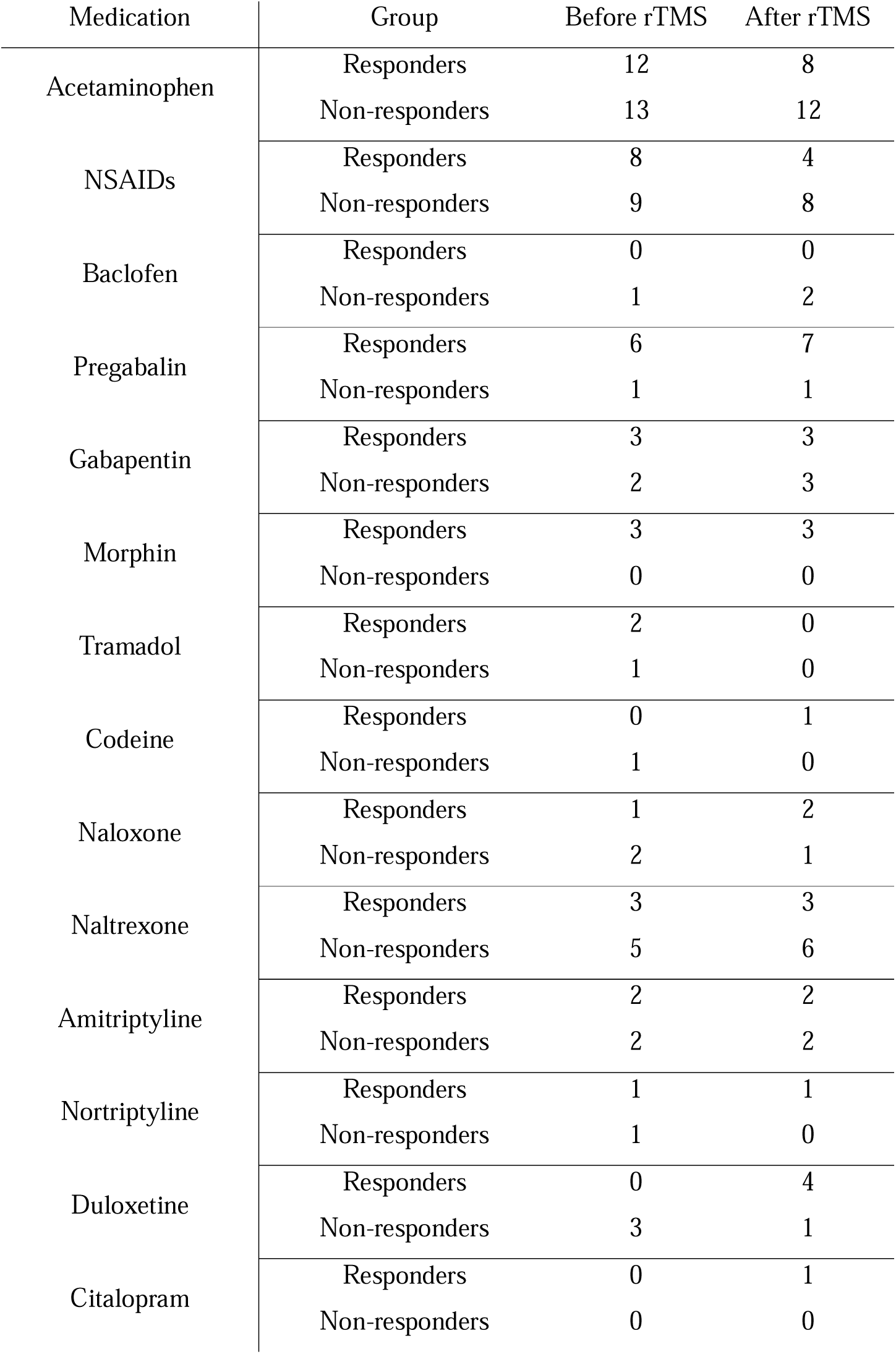
Table showing the number of responders (total n=20) and non-responders (total n=25) using a specific medication before and after rTMS treatment (week 8).

| Medication | Group | Before rTMS | After rTMS |
| --- | --- | --- | --- |
| Acetaminophen | Responders | 12 | 8 |
|  | Non-responders | 13 | 12 |
| NSAIDs | Responders | 8 | 4 |
|  | Non-responders | 9 | 8 |
| Baclofen | Responders | 0 | 0 |
|  | Non-responders | 1 | 2 |
| Pregabalin | Responders | 6 | 7 |
|  | Non-responders | 1 | 1 |
| Gabapentin | Responders | 3 | 3 |
|  | Non-responders | 2 | 3 |
| Morphin | Responders | 3 | 3 |
|  | Non-responders | 0 | 0 |
| Tramadol | Responders | 2 | 0 |
|  | Non-responders | 1 | 0 |
| Codeine | Responders | 0 | 1 |
|  | Non-responders | 1 | 0 |
| Naloxone | Responders | 1 | 2 |
|  | Non-responders | 2 | 1 |
| Naltrexone | Responders | 3 | 3 |
|  | Non-responders | 5 | 6 |
| Amitriptyline | Responders | 2 | 2 |
|  | Non-responders | 2 | 2 |
| Nortriptyline | Responders | 1 | 1 |
|  | Non-responders | 1 | 0 |
| Duloxetine | Responders | 0 | 4 |
|  | Non-responders | 3 | 1 |
| Citalopram | Responders | 0 | 1 |
|  | Non-responders | 0 | 0 |

Among Responders, 6 participants (30%) reported being “Very much improved” (PGIC) and “Much improved,” followed by 9 patients (45%) reporting “Minimally improved” at the end of treatment. However, 5 Responders (25%) reported “No improvement”. Non-responders most commonly reported “Minimally improved” (12 participants, 48%), followed by “No improvement” (11 participants, 44%). One Non-responder (4%) rated “Much improved”, while 1 (4%) rated “Minimally worse”.

### TMS-EEG global and local mean field power

The GMFP did not differ significantly between responders and non-responders (t_43_ = 0.556, P = 0.581; Figure 3A). Moreover, in the electrode cluster around the stimulated area, LMPF also did not differ significantly between groups (t_43_ = 1.845, P = 0.072; Figure 3B).

### TMS-EEG local oscillatory dynamics

ERSP was higher in responders compared with non-responders (Mann–Whitney U = 2.672, P = 0.008; Figure 3C). Similarly, ITC was increased in the Responder group (t_43_ = 2.741, P = 0.010, 95% CI [0.04, 0.24]; Figure 3D).

### Correlation analyses

High levels of local connectivity correlated with post-treatment pain relief. Including all patients, the percentage of pain reduction correlated with alpha-ERSP (Spearman rL=L-0.36, PL=L0.014 - Figure 4A) and alpha-ITC (Spearman rL= −0.33, PL=L0.026 - Figure 4B).

**Figure 4:**
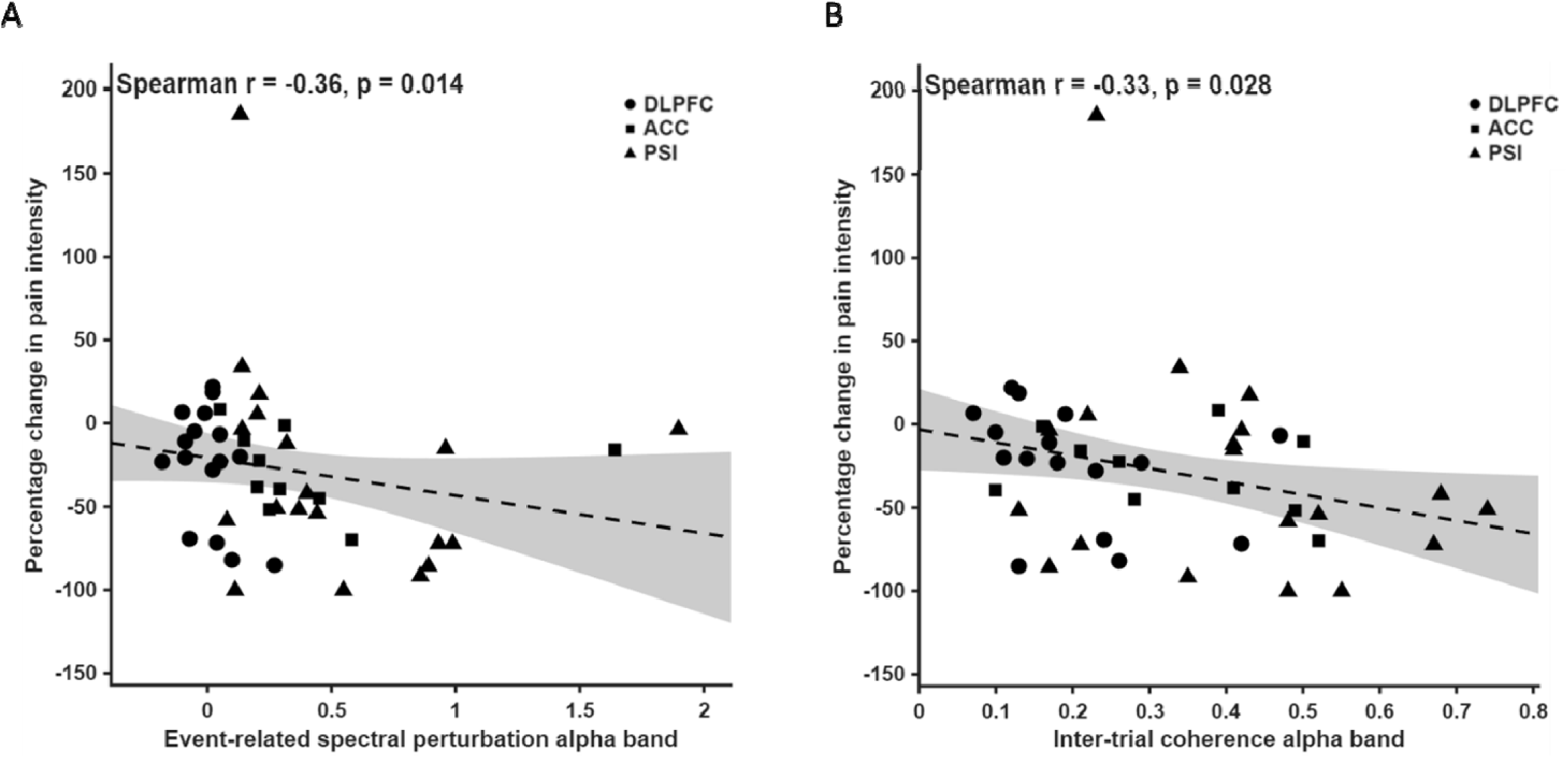
Correlation between percentage change in pain intensity and neurophysiological outcomes. **A)** Event-related spectral perturbation in the alpha band (dB). **B)** Inter-trial coherence in the alpha band (0–1).

## DISCUSSION

The current study showed that responders to non-motor rTMS exhibited higher local TMS-evoked alpha-band oscillatory dynamics before treatment compared with non-responders. This suggests that greater pre-treatment alpha-band power and phase in electrode clusters around the targeted region may reflect a cortical connectivity state more favorable to modulatory neuroplastic changes induced by rTMS, thereby increasing the likelihood of a positive response to non-motor targeting. Identifying such oscillatory signatures could help detect patients who are more likely to benefit from non-motor rTMS interventions for chronic pain and enrich clinical trials by using biomarkers of treatment response.

### Clinical outcomes

In this secondary exploratory analysis, 44% of patients experienced a clinically meaningful pain reduction after rTMS delivered to various non-motor cortical regions. Among these responders, 11 received stimulation to the PSI (58% of all PSI cases), 5 to the ACC (50% of all ACC cases), and 4 to the DLPFC (25% of all DLPFC cases). These results extend previous findings on the clinical effects of rTMS applied to the PSI, ACC, and DLPFC [19,21,37,61].

Regarding the PSI, a previous trial in patients with peripheral neuropathic pain reported that 58% (18 out of 31) achieved a clinically meaningful pain reduction [19], which is comparable to the proportion observed in the current study. In contrast, in a study of 33 patients with central neuropathic pain, no analgesic effect was observed in most patients, although stimulation produced significant increases in heat pain and warm detection thresholds, indicating antinociceptive changes that were dissociated from the clinical efficacy of this approach [16]. Notably, there are important methodological differences among the studies. In the current study and the trial by Dongyang et al. [19] the left or right PSI was stimulated contralateral to the main painful area. In contrast, Galhardoni et al. stimulated only the right PSI, regardless of symptom laterality [16]. This distinction suggests that targeting the PSI receiving the predominant nociceptive input may be critical for achieving analgesic effects. Another important observation is that Galhardoni et al. included only patients with central neuropathic pain [16], while both Dongyang et al. and the current study enrolled individuals with peripheral neuropathic pain [19]. In effect, many responders in the current trial had peripheral neuropathic pain, nociceptive, or nociplastic pain. This may indicate that PSI stimulation is more likely to produce analgesic effects when nociceptive inputs reach a structurally intact spino-thalamo-insular projection [29], consistent with the established role of this region in processing incoming sensory-discriminative pain signals and modulating nociceptive gain.

Regarding the ACC, our findings showed that 50% of patients (5 out of 10) responded to stimulation. Two previous studies have investigated rTMS of ACC in chronic pain, with contrasting results. A study with 16 fibromyalgia patients reported a 43% mean reduction in pain intensity 4 weeks after completing 20 sessions of rTMS to the ACC [21]. In a larger study of 33 patients with central neuropathic pain, no analgesic effect of ACC-rTMS was observed, but the stimulation produced a significant reduction in HADS scores [16]. In our cohort, ACC responders showed a mean HADS reduction of 36 ± 42%, whereas non-responders showed a mean reduction of 5 ± 18%. Together, these findings suggest that ACC stimulation may not provide a clear analgesic effect, but it may influence affective symptoms associated with chronic pain

For the left DLPFC, conflicting results have been reported, with some studies showing an analgesic effect [17,23,37,38,62], while others failed to detect a therapeutic benefit [1]. In the current study, only 25% of patients responded to DLPFC rTMS (4 out of 16), which is comparable to the response rate reported in DLPFC and sham groups in previous trials [1]. Notably, studies demonstrating pain reduction typically targeted specific pain conditions often associated with co-morbid depressive or affective symptoms, such as fibromyalgia [17], migraine [22], and burning mouth syndrome, or with anxiety, as in chronic low back or neck pain [62]. Together, these observations suggest that only a subset of chronic pain patients, particularly those with comorbid major depressive disorder [63], may benefit from DLPFC stimulation. Because our study and other negative ones [1] excluded individuals with these conditions, this may explain the low response rate observed in the DLPFC group.

### TMS-evoked oscillatory dynamics

In the current study, we found a significant increase in alpha-band oscillatory dynamics in responders compared with non-responders. TMS-evoked oscillatory dynamics refer to the rhythmic neural activity induced by TMS, expressed as changes in oscillatory power and phase across different frequency bands relative to pre-stimulus activity [64]. Because TMS provides an external perturbation to the cortex, it enhances the expression of intrinsic oscillatory properties, yielding robust and frequency-specific responses that reflect the local cortical dynamics of the stimulated region [65]. Several previous studies have demonstrated its effectiveness in discriminating between normal and abnormal cortical oscillatory patterns in different pathological conditions, such as strokes [66], Parkinson’s disease [67], unresponsive wakefulness syndrome [68], schizophrenia [69], and major depression [70]. In this study, responders showed higher pre-treatment alpha-band power and phase than non-responders, and this increase correlated with the magnitude of pain relief. In a healthy individual, alpha power reflects active inhibitory gating of sensory or cognitive processing [71], while alpha phase coordinates pulsed inhibition and rhythmic sampling of input [72]. In this context, the elevated alpha power and phase observed in responders may reflect an abnormally high level of synchronization within the probed cortical networks. Thus, increased alpha power and phase in these areas may represent a pathological deviation from their natural oscillatory profile, and rTMS may exert its therapeutic effect by restoring these rhythms toward a more physiological range, consistent with homeostatic plasticity principles [73]. According to these principles, when stimulation is applied to an already hyperactive area, neural circuits tend to down-regulate their activity back toward the physiological state [74]. Consequently, patients with elevated pre-treatment alpha oscillatory activity in non-motor areas may have the appropriate neurophysiological substrate for 10 Hz rTMS to induce a modulatory effect. Interestingly, our previous work on M1 rTMS in patients with chronic pain found that responders exhibited reduced alpha-band power and phase [9]. Taken together, the results of these two studies suggest that effective neuromodulation depends on the momentary state of oscillatory abnormality of the targeted region. While M1 rTMS responders benefited when alpha-band activity was low (i.e., reflecting defective GABA-dependent intracortical inhibition [75]), non-motor rTMS responders benefited when alpha-band activity was overly high (i.e., indicating an abnormally elevated inhibitory state). This suggests that rTMS may be most effective when it modulates target-specific oscillatory deviations, downregulating hyperactive rhythms in non-motor regions or upregulating hypoactive rhythms in M1. Of course, this interpretation is speculative and must endure the test of both human and non-human mechanistic experimental studies.

These findings, along with previous results [9] suggest important clinical implications for personalizing rTMS therapy in chronic pain. The optimal neurophysiological state for effective neuromodulation appears to differ across cortical targets. Such target-specific signatures could guide clinicians in selecting the most appropriate stimulation site for each patient, allowing for a truly personalized neuromodulatory approach to chronic pain management.

### Limitations

Several limitations should be acknowledged. First, this was a secondary exploratory analysis with a relatively small sample size within each stimulation target, which may limit the generalizability of the findings. Nonetheless, most existing TMS–EEG and non-motor rTMS studies are similarly constrained by small samples, and the present results expand current knowledge in this understudied area.

Second, the study included patients with a broad range of chronic pain etiologies, introducing heterogeneity in neurophysiological profiles and treatment responsiveness. However, the observation that patients with diverse etiologies benefited from the intervention suggests that rTMS may be effective beyond neuropathic pain and fibromyalgia, broadening its potential clinical application.

Third, deep TMS inevitably activates not only deeper structures, such as the ACC and PSI, but also the overlying superficial cortex. Thus, while TMS–EEG recordings from these cortical targets may provide valuable biomarkers for non-invasive neuromodulation, theoretical models derived from a technique with limited spatial resolution should be interpreted with caution. It must be kept in mind that the actual targets of the stimulations cannot be fully known. Our navigation system was based on scalp locations/electrodes that were orthogonally positioned relative to the putative cortical areas targeted.

## Conclusion

Using rTMS delivered to non-motor cortical targets produced clinically relevant pain relief in 44% of patients with chronic pain, with response rates varying across PSI, ACC, and DLPFC. Pre-treatment TMS–EEG measures showed that responders had increased alpha-band oscillatory dynamics at the stimulated site, suggesting that the neurophysiological state of the target region plays a key role in determining treatment efficacy. These findings may suggest that rTMS exerts its benefits by modulating region-specific oscillatory abnormalities rather than producing uniform effects.

## Data Availability

All data produced in the present study are available upon reasonable request to the authors

## Acknowledgements

The Center for Neuroplasticity and Pain (CNAP) is supported by the Danish National Research Foundation (DNRF121). The current study is supported by a Novo Nordisk (Grant NNF21OC0072828) and an ERC Horizon Europe Consolidator grant (PersoNINpain 101087925). TGN receives funding from the Lundbeck Foundation (R441-2023-232). All data are available upon reasonable request to the corresponding author.

## Author contribution

**Enrico De Martino:** Conceptualization, Methodology, Formal analysis, Investigation, Data curation, Visualization, Writing- Original draft preparation; Project administration; **Margit Midtgaard Bach**: Investigation, Data curation, Writing - Review & Editing; **Bruno Andry Nascimento Couto**: Software, Formal analysis, Data Curation, Visualization, Writing - Review & Editing; **Anne Jakobsen**: Investigation, Writing - Review & Editing; **Stian Ingemann-Molden**: Investigation, Writing - Review & Editing; **Adenauer Girardi Casali**: Software, Visualization, Writing - Review & Editing; **Thomas Graven-Nielsen**: Conceptualization, Methodology, Writing - Review & Editing; Funding acquisition **Daniel Ciampi de Andrade**: Conceptualization, Methodology, Visualization, Writing - Review & Editing; Project administration, Funding acquisition.

